# Underlying co morbidity reveals unique immune signatures in Type II diabetes patients infected with SARS-CoV2

**DOI:** 10.1101/2021.12.03.21267282

**Authors:** Soumya Sengupta, Gargee Bhattacharya, Sanchari Chatterjee, Ankita Datey, Shubham K Shaw, Sandhya Suranjika, Paritosh Nath, Prakash K Barik, Punit Prasad, Soma Chattopadhyay, Rajeeb K Swain, Ajay Parida, Satish Devadas

**Author notes:** These authors contributed equally (Soumya Sengupta, Gargee Bhattacharya). Address of Corresponding authors: Dr Rajeeb K Swain (RS) Cancer Biology, Institute of Life Sciences, (Autonomous Institute of Dept of Biotechnology, Govt. of India), Nalco Square, Bhubaneswar-751023. Odisha, India, Dr. Ajay Parida (AP), Plant and Microbial Biotechnology, Institute of Life Sciences, (Autonomous Institute of Dept of Biotechnology, Govt. of India), Nalco Square, Bhubaneswar-751023. Odisha, India, Dr. Satish Devadas (SD), Infectious Disease Biology, Institute of Life Sciences Institute of Life Sciences, (Autonomous Institute of Dept of Biotechnology, Govt. of India), Nalco Square, Bhubaneswar-751023. Odisha, India.

## Abstract

**Background:** SARS-CoV2 infection in patients with comorbidities, particularly T2DM has been a major challenge globally. Here, we did whole blood immunophenotyping along with plasma cytokine, chemokine, antibody isotyping and viral load determination from oropharyngeal swab to understand the immune pathology in the T2DM patients infected with SARS-CoV2.

**Methods:** Blood samples from 25 Covid-19 positive patients having T2DM, 10 Covid-19 positive patients not having T2DM and 10 Covid-19 negative, non-diabetic healthy controls were assessed for various immune cells by analyzing for their signature surface proteins in mass cytometry. Circulating cytokines, chemokines and antibody isotypes were determined from plasma. Viral copy number was determined from oropharyngeal swabs. All our representative data corroborated with laboratory findings.

**Results:** Our observations encompass T2DM patients having elevated levels of both type I and type II cytokines and higher levels of circulating IgA, IgM, IgG1 and IgG2 as compared to NDM and healthy volunteers. They also displayed higher percentages of granulocytes, mDCs, plasmablasts, Th2-like cells, CD4^+^ EM cells, CD8^+^ TE cells as compared to healthy volunteers. T2DM patients also displayed lower percentages of pDCs, lymphocytes, CD8^+^ TE cells, CD4^+^, CD8^+^ EM.

**Conclusion:** Our study demonstrated that patients with T2DM displayed higher inflammatory markers and a dysregulated anti-viral and anti-inflammatory response when compared to NDM and healthy controls.

**Contribution to the field:** Covid-19 infection in people with comorbidities, particularly T2DM has been a cause of mortality in several nations and they represent an extremely vulnerable population to Covid-19. This study is one of the most comprehensive study from India, to understand the interplay between immune response and viremia occurring in these T2DM patients infected with SARS-CoV2 and will help in designing public health response and vaccination priorities.

## Introduction

Since the outbreak of the novel coronavirus disease (Covid-19) in late 2019, India has recorded over 34 million cases and 4,54,745 deaths as of 25^th^ October, 2021 and now has the second highest number of cases in the world, after the United States of America. Essentially, SARS-Cov2 is a betacoronavirus, belonging to Coronaviridae family and is closely related to SARS and MERS, which were responsible for earlier disease outbreaks in 2003 and 2011 respectively(1). Broadly, Covid-19 cases were and are still classified into symptomatic and asymptomatic, based on the presence or absence of symptoms and eventual severity based on immune response and/or its failure. With respect to the symptoms of Covid-19 infection, symptomatic patients have fever, dry cough, shortness of breath, acute respiratory distress, loss of taste and smell and in certain cases diarrhoea (2, 3). Based on the intensity of these symptoms, the patients are classified into mild and severe, during diagnosis. Prognosis of the severe patients is critical. However, the major challenge arises from the huge population of asymptomatic cases, as they are responsible for the undetected spread of infection.

In general, disease severity in Covid-19 is associated with lymphopenia, cytokine storm, blood coagulation, drop in pO_2_ levels, etc. (4, 5). However, with respect to the Indian population, these clinical parameters and the associated immune response were not definitely present in most Covid-19 infected patients. Interestingly, one of the major attributes of Covid-19 in Indian population, especially in the first wave, was quick recovery, but the underlying immunological mechanism was and is yet to be understood. However, similar to the first wave, 70% of India’s mortality in Covid-19 in the second wave is still attributed to comorbidities, specifically type II diabetes (T2DM). Although vaccinations drive initiated by the government aims to protect the population in general, the overwhelming number of these patients poses a major challenge to recovery and recuperation of the individual. In that respect, co-relation between comorbidities including T2DM and viral load, T2DM and glucose levels, etc. in the patient can play a major role in dictating the immune response and eventual outcome of Covid-19 infection. Therefore, it is crucial to understand the interplay of these factors along with immunological parameters to give us a comprehensive idea about the status of T2DM patients having Covid-19.

Immune response can be divided into three types namely, type I (antiviral), type II (anti helminthic), type III (antifungal). Type I response mainly constitutes of T-bet and Interferon gamma (IFN-γ) mediated response, which is against intracellular pathogens, including viruses. On the other hand, type II response is mediated by GATA-3 and is mainly against helminthic worms, which are carried out by effector molecules such as IL-4, IL-9, and IgE. The third type of response (type III response) is mediated by ROR-γt and effector molecules such as IL-17A, IL-17F, 1L-22 etc. are responsible for controlling fungal infections (6, 7). Longitudinal analysis has shown an immune dysregulation in Covid-19 patients and people with T2DM contracting Covid-19 Infection had higher innate immune cells, lower T lymphocytes, a sustained increase in antiviral, anti-fungal response and higher type 2 response such as IL-5, IL-13, IgE and eosinophils (8).

Altogether, the immunological response in Covid-19 patients with co-morbidity is an evolving area of study and these patients represents higher vulnerability in comparison to the ones without any comorbidity or other comorbidities. Our study aims to elucidate the differences between T2DM patients having Covid-19 as compared to those devoid of T2DM, based on multiple parameters such as viral load, cytokine *milieu* and immune cells, which will give us a comprehensive idea about the role played by T2DM in Covid-19 pathogenesis.

## Materials and Methods

### Population and Samples

A total of 25 samples with T2DM and 10 samples without T2DM (NDM) (Non-diabetes mellitus) were collected. These samples were collected from hospital within 4 days of patient’s admission. Type II diabetes in these patients was confirmed by previous clinical history and for this particular study was assessed by fasting plasma glucose level and glycated haemoglobin and other clinical and biochemical parameters as requested by their consultant medical practitioner. Amongst these, 22 patients with T2DM had hypertension. The T2DM or NDM patients neither reported nor were assessed for other comorbidities during the infection. The detailed laboratory findings are given in Table 1. All the T2DM patients had severe symptoms, as assessed by the clinician. The NDM patients either had mild symptoms or were asymptomatic. Additionally, 10 healthy volunteers who tested negative for Covid-19 were included in the study to provide basal and or steady state biochemical and cytokine profile. Briefly, 5ml blood was collected in BD Vacutainer EDTA tubes (BD 367863) and 270 μl of whole blood from 10 T2DM patients, 5 NDM patients and 5 healthy volunteers was used for deep immune profiling by mass cytometry. The tubes were spun at 1600g for 20 minutes at room temperature. Plasma was then collected and stored in −80 °C till further analysis. The details of all the reagents and software used are given in the supplementary table 1 (S1).

**Table 1-.** Demographic details and laboratory findings of T2DM and NDM patients infected with SARS-CoV2.

|  | T2DM<br>n=25 | NDM<br>n=10 | P VALUE |
| --- | --- | --- | --- |
| Age, median (IQR), years | 46 (36-59) | 49(40.25-55.50) | N. A |
| Sex, male/female, n (%) | 17 (68%)/8(32%) | 7 (70%)/3(10%) | N. A |
| Haemoglobin, gm/dl median (IQR) | 12.2 (11.35-13.60) | 14.25 (13.7-14.9) | 0.0475 |
| Platelets 10<sup>3</sup>/μL median (IQR) | 193 (154.5-235) | 165 (159.5-197.5) | 0.4027 |
| White Blood Cells 10<sup>3</sup>/μL, median (IQR) | 6.1 (5.00-57.65) | 5.8 (5.2-5.575) | 0.4172 |
| Neutrophils %, median (IQR) | 75 (65.5-84) | 55 (44.75-62) | 0.0004 |
| Lymphocytes%, median (IQR) | 22 (15.5-29.75) | 36 (26.75-39.75) | 0.0349 |
| Eosinophils %, median (IQR) | 0 | 3 (3-4.5) | <0.0001 |
| Monocytes%, median (IQR) | 7 (5-9) | 9 (8.5-10) | 0.0373 |
| D-dimer μg/mL, median (IQR) | 0.45 (0.29-1.61) | 0.19 (0.15-0.25) | 0.0048 |
| Ferritin ng/mL, median (IQR) | 224.7 (145.9-464.1) | 115 (110-289.5) | 0.4975 |
| Fasting Plasma Glucose mg/dL, median (IQR) | 215 (142.72-244) | 95.5 (88-101) | <0.0001 |
| Hb1AC %, median (IQR) | 6.9 (6-9) | N. T | N. A |
| CRP mg/mL, median (IQR) | 69.62 (34.95-211) | 13.28 (4.44-18.65) | 0.0013 |
| Serum SGOT U/L, median (IQR) | 46.9 (30.2-64.6) | 38 (30.75-43.25) | 0.4432 |
| Serum SGPT U/L, median (IQR) | 27.6 (20-42) | 40.5 (24.5-76.5) | 0.1335 |

**Hb1AC- Glycated haemoglobin, CRP- C-Reactive Protein, Serum SGOT-serum glutamic-oxaloacetic transaminase, Serum SGPT- serum glutamic-pyruvic transaminase, N.T-Not Tested, N.A-Not Applicable.**

### SARS-CoV2 Viral load detection in respiratory specimen and plasma

300 μl each of oropharyngeal swab and plasma were taken for viral RNA extraction using TAN Bead Maelstrom 4800 as per manufacturer’s instructions. The extracted viral RNA was stored at-80°C until further use (9).

### qRT-PCR

The qRT-PCR was performed using 5μl of the extracted RNA from samples using the TRUPCR SARS-CoV-2 RT qPCR Kit V-2.0. The human RNase P served as an internal control whereas envelope (E) and nucleocapsid (N) genes were targeted for SARS-CoV-2 amplification (9).

### Viral copy number determination

The viral copy number was determined for the above-mentioned samples by generating the standard curve of SARS CoV-2 N (nucleocapsid) gene. The N gene was cloned into pBiEx vector and 10-fold serial dilutions of the plasmid were done to obtain the standard curve. The percentage of copy number/ml was calculated from the corresponding Ct values of all the samples. For obtaining Ct values, cDNA was prepared from the extracted RNA using random hexamers by TAKARA primescript 1st strand cDNA synthesis kit (Kusatsu, Japan). The cDNA was subjected to qPCR (Mesagreen SYBR Green-No ROX, Eurogentec, Belgium) using nucleocapsid gene specific primers. (FP: GTAACACAAGCTTTCGGCAG and RP: GTGTGACTTCCATGCCAATG) (9).

### Plasma Cytokine and Chemokine detection assay

Neat plasma from Covid-19 and controls was used to measure 41 cytokines and chemokines using human Milliplex map cytokine assay kit (Millipore, Billerica, MA, USA). The samples were acquired in a Bio-Plex 200 system (Bio-Rad, Hercules, CA, USA) and cytokine concentrations were calculated using Bio-Plex manager software with a five-parameter (5PL) curve-fitting algorithm applied for standard curve calculation (10).

### Isotyping of circulating antibody from blood

For analysis of the isotype composition of antibodies in circulation, plasma of T2DM, NDM patients and healthy volunteers, were analysed by ProcartaPlex Human Antibody Isotyping Panels (Cat. No EPX070-10818-901, Invitrogen, Vienna, Austria), based on the manufacturer’s instructions. Briefly, antibody-coated magnetic bead mixtures were incubated with 25μl of assay buffer, kit standards or diluted plasma (1:20000) samples in a ProcartaPlex 96-wells plate at room temperature for 1 hour. Detection antibodies (25 μl) were then added and the plates were incubated on an orbital shaker at 500 rpm for 30 min. Next, the wells were incubated with 50μl of diluted Streptavidin-Phycoerythrin for 30 min. Plates were then washed using a hand-held magnetic plate washer. All incubations were performed at room temperature in the dark. Afterwards, samples were suspended in 120 μl reading buffer. The samples were acquired in a Bio-Plex 200 system (Bio-Rad, Hercules, CA, USA) and cytokine concentrations were calculated using Bio-Plex manager software with a five-parameter curve-fitting algorithm (5PL) applied for standard curve calculation (11).

### Whole blood Immunophenotyping by Mass Cytometry

For immunophenotyping, 270μl of whole blood was collected, added to pre-coated Maxpar Direct Immune Profiling tubes and incubated for 30 minutes in room temperature. For lysis, 250μl of 1x BD FACs Lyse was added, followed by 10 min incubation. There were two consecutive washes with Maxpar water and Cell staining buffer. The cells were then fixed with 4% formaldehyde and incubated for 10 minutes followed by washing at 800g for 5 minutes. Eventually, the cells were suspended in 1ml Iridium solution and stored in −80 °C till acquisition. For acquisition, the cells were thawed, washed with Cell Staining buffer and Cell Acquisition Solution. The cell density was adjusted to 1 million cells/ml in Cell acquisition Solution with 0.1% EQ Beads and acquired in Helios Mass cytometer. FCS files were then normalised with CyTOF software V.7 (Fluidigm) and then exported and analysed by FlowJo software V10.7 (BD Biosciences)(12, 13).

### Statistics

Statistical analysis was performed using the GraphPad Prism software, version 8.0.1. Data was presented as Mean ± Standard Deviation of Mean (SEM). Non-parametric Kruskal Wallis Test with post hoc Dunn’s multiple comparison test was used to compare the levels of cytokines, antibodies and percentage of different immune cells among the three groups. Mann-Whitney U test was used to compare laboratory parameters between T2D and NDM patients. *P* values less than 0.05 were considered significant (\**P* < 0.05, \*\**P* < 0.01, \*\*\**P* < 0.001, \*\*\*\**P* < 0.0001).

### High dimensional analysis

For high dimensional analysis, t-SNE analysis was performed using FlowJo V.10.7 (BD Biosciences). The gating for analysis was done on live, intact, CD45^high^CD66b^low^ lymphocytes. Amongst a total of 100,000 events, 5000 events in the lymphocyte population were used per sample for this analysis. 5 samples of T2D and of Healthy controls were used to compute t-SNE and a total of 25000 events per group were concatenated and exported for t-SNE analysis. The default parameters in the software, iterations-1000, perplexity-30, eta-675, KNN algorithm-Exact (vantage point tree), gradient logarithm-Barnes-Hut were used to compute the t-SNE plot.

## Data Availability

The human clinical data is stored at KIMS Hospital, samples are stored at ILS Biorepository, Ethical clearance with the corresponding authors and on request for this, ethical, legal, and privacy consent will be requested from both the doctor and patient to share data.

## Ethics Statement

The studies involving human participants were reviewed and approved by the Institutional Human Ethics Committee, Institute of Life Sciences. The Institutional Ethics Committee (IEC)/ Institutional Review Board (IRB) reference number is 106/HEC/2021. Written consent informed to participate in this study was provided by the participants’ spouse/next of kin.

## Author Contributions

Experimental Design and Conceptualization – SD, SS and GB

Sample Collection and processing-RS, PP, SaS, SS, GB, SaC, PB and PN

Mass Cytometry Experiments-SS, GB and SKS

Chemokine and Cytokine Multiplexing-SS, SaC and GB

Antibody Isotyping-SS, SaC and AD

RT-PCR and Virological assays-SaC, AD and SC

Data analysis-SS, GB, SaC, AD, SC and SD

Manuscript Drafting-SS, GB, SaC, SC and SD

Manuscript Editing and Review-GB, SS, RS, PP, AP, SC and SD

## Acknowledgements

We would like to acknowledge ILS Biorepository for providing the SARS-CoV2 samples. We would like to thank Dr. Gajendra Motiram Jogdand for valuable suggestions in multiplexing. We would also like to thank Mr. Deepak Singh, Mr. Satyajit Behera and Ms. Jyotsna Priyadarshini for data entry.

## Results

### Viral load does not corroborate with inflammation in T2DM

The study was executed on NDM and T2DM Covid-19 patients, who had been detected and admitted to a local tertiary COVID hospital. To examine for viral load, oropharyngeal (OP) samples were collected from various patients as per established protocol and processed in the BSL3 facility at ILS, Bhubaneswar. In most of the NDM and T2DM patients, there was no significant difference in the viral copy number or ΔΔ Ct values of OP samples at the time of sampling **(Fig. 1A and B)**. This suggests that the viral load in OP samples does not and did not correlate with disease severity in this population of study. This probably indicates that the viral load present in the host does not dictate the severity of Covid-19 infection.

**Fig 1.**
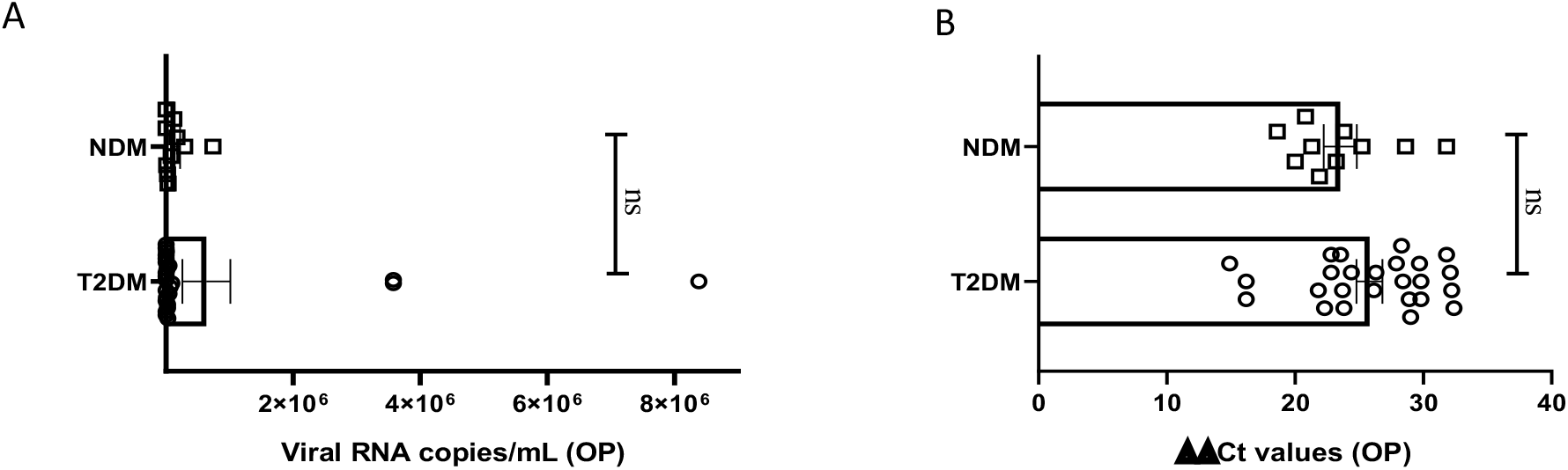
Analysis of Viral load in Oropharyngeal samples in Covid-19 patients. **(A)** Bar diagram depicting the viral copy number of Oropharyngeal (OP) samples collected from T2DM(n=25) and NDM(n=10) COVID-19 patients.(B) Bar diagram representing the **ΔΔ** CT of OP samples of NDM and T2DM COVID-19 patients. The Mann-Whitney (non parametric, two tailed) test was performed. ns, not significant. All error bars were SEM.

### Altered cytokine and chemokine profiles in Covid-19 patients with Type II Diabetes

To investigate the status of systemic inflammation concurrent with SARS-Cov2 infection, we did multiplexing analysis of 41 cytokines and chemokines from the plasma of T2DM and NDM patients to delineate the altered immune microenvironment in these patients. For comparison and for basal level expression of these cytokines and chemokines, we added ten healthy volunteers and segregated the proteins into significant, moderate and mild based on the level of their expression in T2DM as compared to NDM and healthy volunteers. Accordingly, the levels of IL-6, TNF-α, G-CSF, GM-CSF, IFN-α2, IL-10, VEGF, IL-1Rα, IL-12p40, IL-15, IL-1α, MIP-1β, were found to be significantly elevated (p<0.0001, Kruskal Wallis Test), indicating the simultaneous release of both Type I and Type II cytokines in severity (**Fig. 2A)**. In contrast, there was moderate elevation in the levels of IFN-γ, EGF, IL-8, indicative of the ongoing antiviral reaction in these patients (**Fig. 2B)**. Amongst the slightly elevated ones, IP-10 (p=0.0365), IL-9, IL-4 (p=0.0146) showed higher significance **(Fig. 2C)**. Although IP-10 has been used as a real time marker for mortality in Covid-19 patients, we did not do a longitudinal patient tracking, therefore cannot ascertain its role in mortality. In addition, IL-4 and IL-9 which are both type II cytokines have also been linked to severity of Covid-19 patients. Although our study could not find any significant difference in levels of type III signature cytokines such as IL-17 and IL-1β, the presence of type I and type II immune signatures were overlapping in multiple patients (data not shown). However T2DM patients showed elevation of both type I and type II cytokines indicating a dysregulated immune response, prominent in these patients as earlier reported by Lucas et al **(Fig. 2)**.

**Fig 2.**
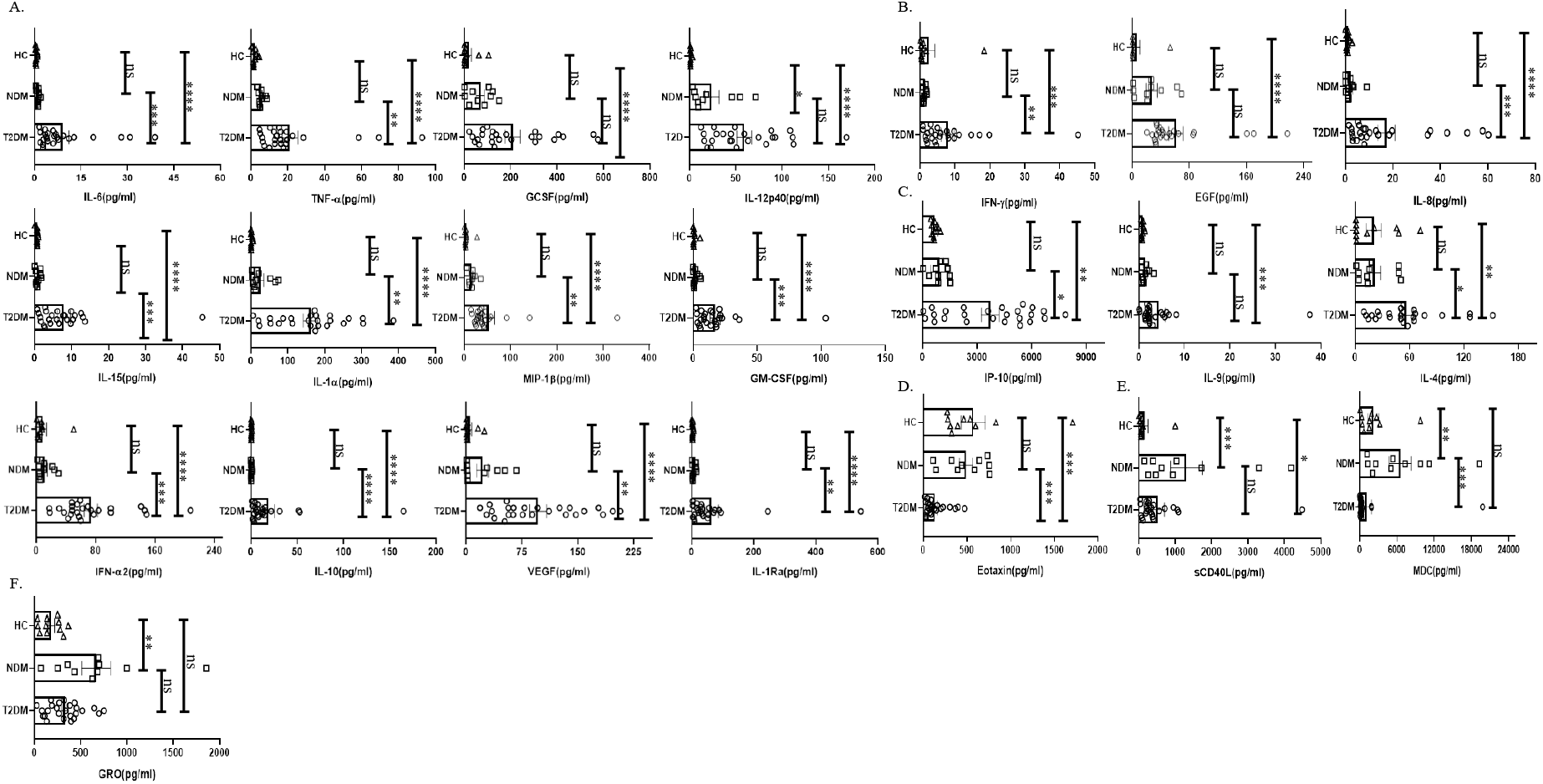
Analysis of the cytokines and chemokines in T2DM, NDM patients infected with SARS-CoV2. The bar diagrams representing cytokines and chemokines which were evaluated from COVID-19 positive plasma samples of T2DM(n=25), NDM(n=10) and Covid-19 negative and non-diabetic healthy controls(n=10).**(A)**IL-6.TNF-α,GCSF,IL-12p40, IL-15,IL-1α,MIP-1β,GMCSF,IFN-α2,IL-10,VEGF and IL-Rα were highly elevated in T2DM patients when compared with healthy controls and NDM patients. **(B)** IFN-γ, EGF and IL-8 were moderately elevated in T2DM patients when compared to healthy controls and NDM.**(C)** IP-10, IL-9 and IL-4 were slightly elevated in T2DM patients when compared to healthy controls and NDM**(D)** Eotaxin was moderately elevated in healthy controls and NDM when compared to T2DM patients.**(E)** sCD40L was elevated in both NDM and T2DM when compared to healthy controls, MDC was elevated in NDM patients when compared to T2DM patients.**(F)**GRO was elevated in NDM patients when compared to healthy controls. The Kruskal Wallis Test (non parametric) with post hoc Dunn’s multiple comparison test was performed p<0.05 was considered statistically significant (*), p<0.01 was considered to be very significant (**), *P* < 0.001 was considered to highly significant(***), *P* < 0.0001 was considered extremely significant(****) ns, not significant. All error bars were SEM.

Interestingly, the levels of chemokines as opposed to cytokines, were higher in NDM and healthy volunteers as opposed to T2DM patients. For example, the levels of Eotaxin were found to be lower in T2DM than in NDM patients and healthy controls (**Fig 2D)**. In general, Eotaxin is associated with chemoattraction of eosinophils, basophils and neutrophils. Previous studies have reported its increase with severity but in our study, the population indicated a reverse trend; decreasing Eotaxin levels in the T2DM patients. This can be indicative of an impaired healing process as previous reports have suggested a positive correlation between increasing number of eosinophils and healing. In addition, sCD40L was elevated in NDM patients as compared to T2DM and healthy volunteers **(Fig. 2E)**. Since sCD40L has been associated with immunosuppression, it is indicative of an anti-inflammatory immune response in NDM patients, which is clearly absent in T2DM cases. Similarly, the levels of MDC (macrophage derived chemokine) were also higher in NDM as compared to T2DM patients, demonstrating its potential role in immune suppression **(Fig. 2E)**. In addition, GRO levels were higher in both NDM and T2DM patients as compared to healthy controls **(Fig. 2F)**. As GRO serves as a chemoattractant for neutrophils, it is indirectly indicative of the status of underlying inflammation in NDM and T2DM patients. Collectively, our cytokine and chemokine profile of T2DM patients showed an increasing trend in the inflammatory cytokine panel and augmented levels of chemokines that were primarily responsible for repair and angiogenesis.

### Circulating antibody isotype profiling showed a skewed inflammatory profile in Covid-19 patients with Type II Diabetes

Various antibody isotypes correlate with the clinical phenotype of an infection and is therefore an important indicator of the type of immune response. Since cytokine *milieu* in T2DM and NDM patients was skewed, we wanted to define the circulating antibody isotypes in these patients as opposed to healthy controls. We did Luminex based multi plex assay from their plasma and quantified the amount of IgA, IgM, IgE, IgG1, IgG2, IgG3 and IgG4. Here, we found IgG1 and IgG2 to be significantly elevated in T2DM patients as compared to healthy controls (p=<0.0001) and NDM patients (p=0.0024 for IgG1 and p=0.0033 for IgG2), correlating with the higher levels of type I cytokines in these patients. IgE, being a hallmark of type II immune response was also elevated in T2DM as compared to NDM (p=0.0453) and healthy control (p=0.0347). Alongside, IgA being associated with mucosal immunity was also higher in T2DM patients as compared to NDM (p=0.0012) and healthy controls (p=0.0140). As IgM is the first antibody isotype to appear during the course of infection, accordingly we found its levels to be higher in T2DM patients as compared to NDM (p=0.0178) and healthy controls (p=0.0017) **(Fig. 3)**. In summary, the antibody isotype profile was concurrent to the cytokine levels, thus confirming and furthering the status of disease severity in Covid-19 patients.

**Fig 3.**
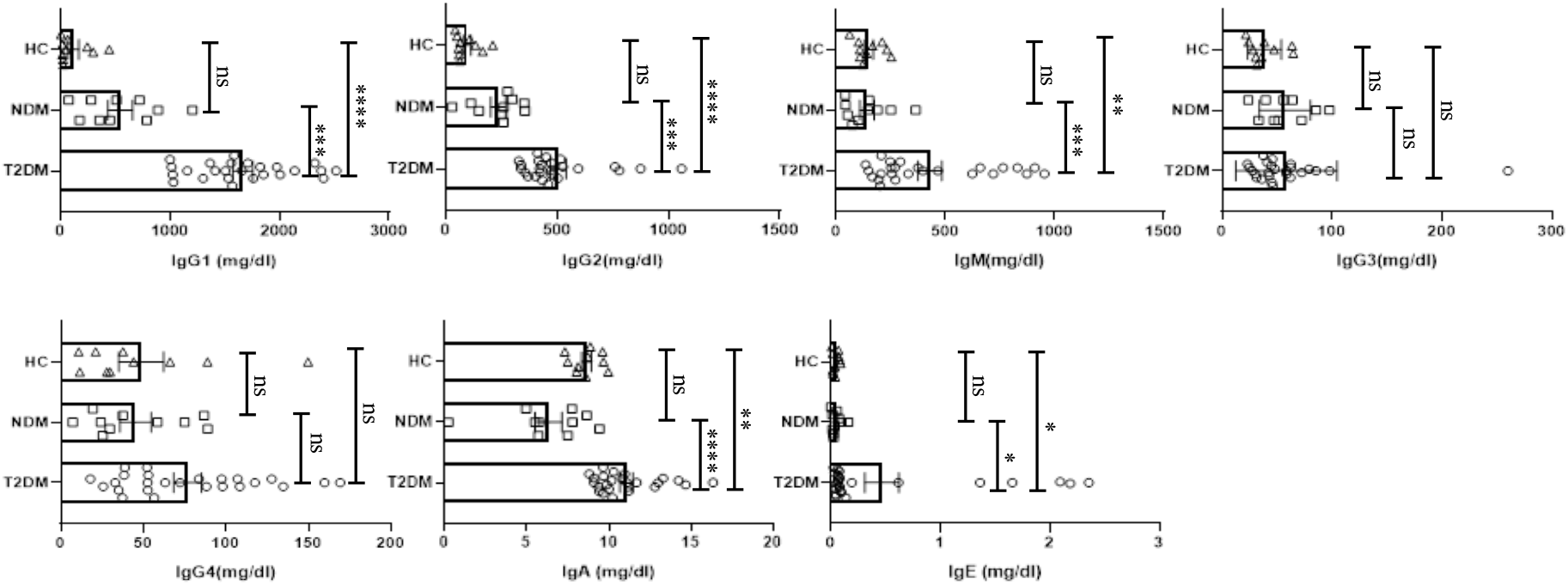
Analysis of the circulating antibodies in T2DM, NDM patients infected with SARS-CoV2. The bar diagram representing circulating antibody isotypes (IgA, IgM, IgG1, IgG2, IgG3,IgG4 and IgE) which were evaluated from COVID-19 positive plasma samples of T2DM(n=25), NDM(n=10) and Covid-19 negative and non-diabetic healthy controls(n=10). The Kruskal Wallis Test (non parametric) withwith post hoc Dunn’s multiple comparison test was performed p<0.05 was considered statistically significant (*), p<0.01 was considered to be very significant (**), *P* < 0.001 was considered to highly significant(***), *P* < 0.0001 was considered extremely significant(****) ns, not significant. All error bars were SEM.

### Dysregulated innate immune system in Covid-19 patients with Type II Diabetes

With the cytokine multiplexing and antibody isotyping indicating an altered immune response, our next objective was to delineate the alteration in the cells of innate and adaptive immune system during Covid-19. Based on the surface markers analysed, we found T2DM patients showing significant increase in *CD45*^*low*^*CD66b*^*high*^ granulocytes as compared to healthy controls (p=0.0074), which is concurrent with the increase in the levels of IL-8 **(Fig. 4A)**. The bulk population of these granulocytes were neutrophils, though we did not find any significant difference between the levels of neutrophils in T2DM, NDM and healthy controls (data not shown). Also, dendritic cell populations were variable, with their numbers decreasing in T2DM as compared to NDM and healthy controls **(Fig. 4B)**. This was in contrast to the increase in IFN-α2 levels, indicating towards dysregulated dendritic cell population in T2DM patients **(Fig. 2A)**. Within the dendritic cell population, plasmacytoid dendritic cells (pDCs) decreased in T2DM patients as compared to NDM and healthy controls **(Fig. 4B)**. However, percentage of myeloid dendritic cells (mDCs) increased in T2DM patients as compared to NDM and healthy controls **(Fig. 4C)**. The percentage of basophils was also reduced in T2DM patients as compared to healthy volunteers, which was correlating with the decrease in levels of chemokine, Eotaxin (CCL11) **(Fig. 4D)**. Although, we did not observe any variation between T2DM, NDM and healthy controls in the populations of gamma delta cells and natural killer cells there was reduction in the percentage of MAIT/NKT cells in T2DM patients as compared to healthy and NDM patients **(Fig. 4E)**. As reported earlier (data not shown), there were no changes in the overall or subsets of monocytes and natural killer cells. Altogether, alterations in the innate immune cell population is suggestive of an aberrant immune response incapable of controlling the infection.

**Fig 4.**
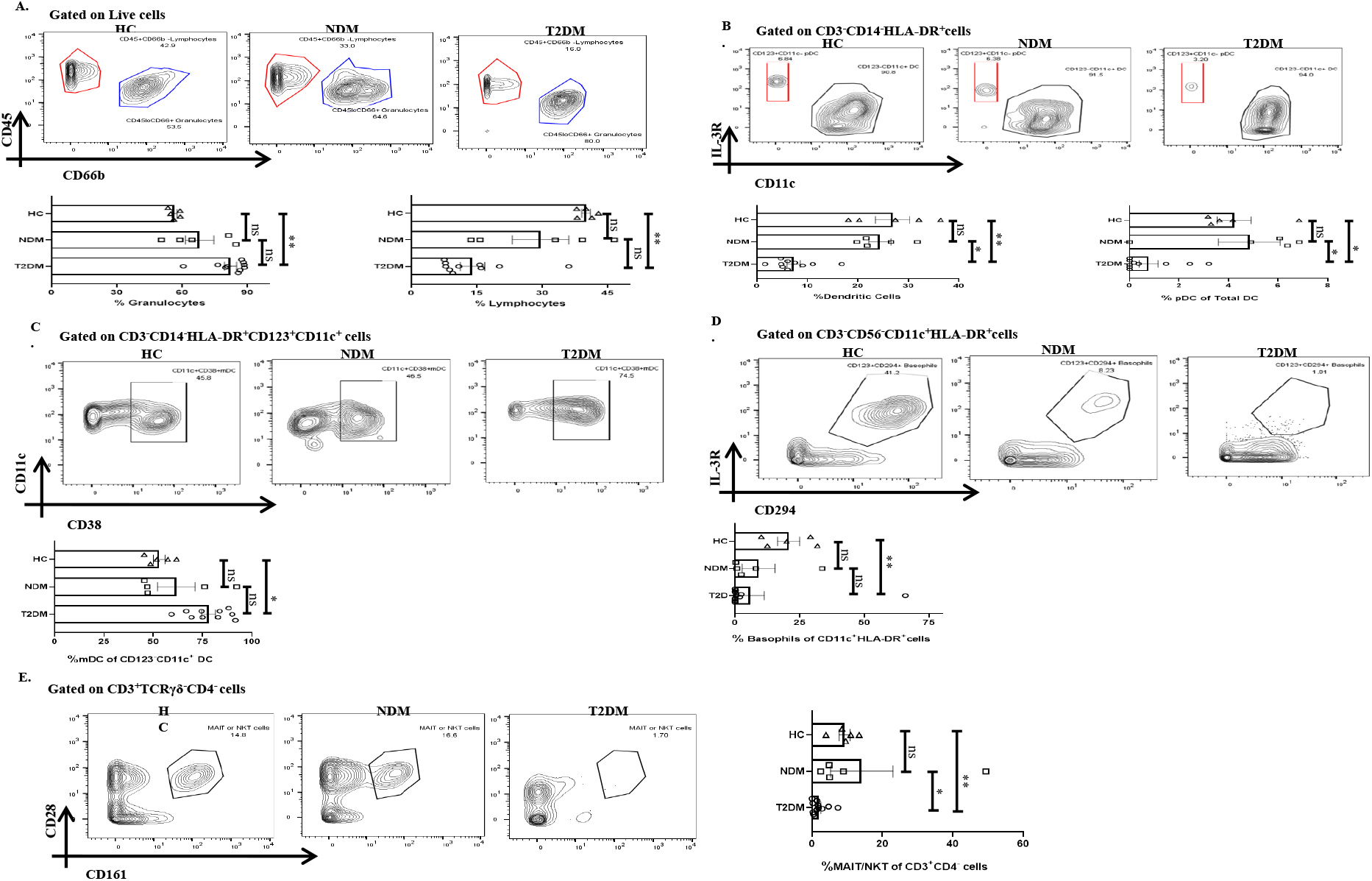
Innate Immune Cells in T2DM and NDM patients. Whole blood from Covid-19 positive T2DM(n=10), NDM(n=5) and Covid-19 negative and non-diabetic healthy controls (n=5)was stained heavy metal tagged antibody and analyzed in mass cytometer. **(A)**Representative Gating strategy to identify CD45^+^CD66b^low^ lymphocytes and CD45^-^CD66b^high^ Granulocytes. Statistical analysis of percentage of Granulocytes and lymphocytes.**(B)** Gating strategy for CD123^+^CD11c^-^ pDCs. Statistical analysis for the frequency of total DCs and pDCs.**(C**) Gating strategy for CD11c^+^CD38^+^ mDCs.Statistical analysis for the percentage of mDCs.**(D)**Gating strategy for CD123^+^CD294^+^ Basophils. Statistical analysis for the frequency of basophils.**(E)** Gating strategy for MAIT/NKT cells.Statistical analysis for the frequency of MAIT/NKT cells.). The Kruskal Wallis Test (non parametric) withwith post hoc Dunn’s multiple comparison test was performed p<0.05 was considered statistically significant (*), p<0.01 was considered to be very significant (**), *P* < 0.001 was considered to highly significant(***), *P* < 0.0001 was considered extremely significant(****) ns, not significant. All error bars were SEM.

### Heterogenicity in Lymphocyte population in Covid-19 patients with Type II Diabetes, NDM and healthy controls

Lymphocytes are instrumental in fighting infections and earlier reports indicated towards its decreasing trend in T2DM patients. There was a moderate decrease in the percentage of lymphocyte in T2DM as compared as to the healthy controls **(Fig. 4A)**. Amongst lymphocytes, percentage of total B cell population was expanding in T2DM patients as compared to healthy controls **(Fig. 5A)** and was is in direct correlation with increased IgG1 and IgG2 in the former **(Fig. 3)**. Further analyses revealed a significant increase in plasmablast population in T2DM patients as compared to healthy controls. Additionally, we found a significant decrease in the percentage of CD3^+^ T cells in T2DM patients as compared to healthy controls suggesting lymphopenia associated with increased severity **(Fig. 5B)**. Within the T cell population, there was an increase in the percentage CD4^+^ T cells (statistically insignificant) but decrease of CD8^+^ T cells **(Fig. 5B)**. Further, in depth analysis of CD4^+^ T cells revealed that T2DM patients had an increase of the Th2 subtype as compared to NDM or healthy volunteers **(Fig. 5C)**. This can be correlated with the increased levels of IL-4 and IL-9, found in the plasma of these patients **(Fig. 2C)**. However, Th1, Th17, Treg and Tfh cells did not show any statistically significant difference, which can be attributed to the time of sampling or be a consequence of dysregulated immune response. There was also no significant difference in Naïve CD4^+^ T cells or Central memory CD4^+^ T cells within the groups. But terminal effector (TE) cells were decreased and effector memory (EM) cells were increased in T2DM patients as compared to healthy controls **(Fig. 5D**). This was in contrast to the CD8^+^ compartment, where TE cells were increased in T2DM patients as compared to NDM but there was no difference between T2DM and healthy controls suggesting a robust response in NDM patients as compared to T2DM and a certain degree of cross-reactive reaction occurring in these T2DM patients **(Fig. 5E)**. Similarly, there was significant decrease of EM cells in T2DM as compared to NDM while no difference was observed in healthy controls when compared to T2DM or NDM. Collectively, a bias was found towards terminal effector CD8^+^ T cells in T cell immune response which was suggestive of an effective ongoing anti-viral response.

**Fig 5.**
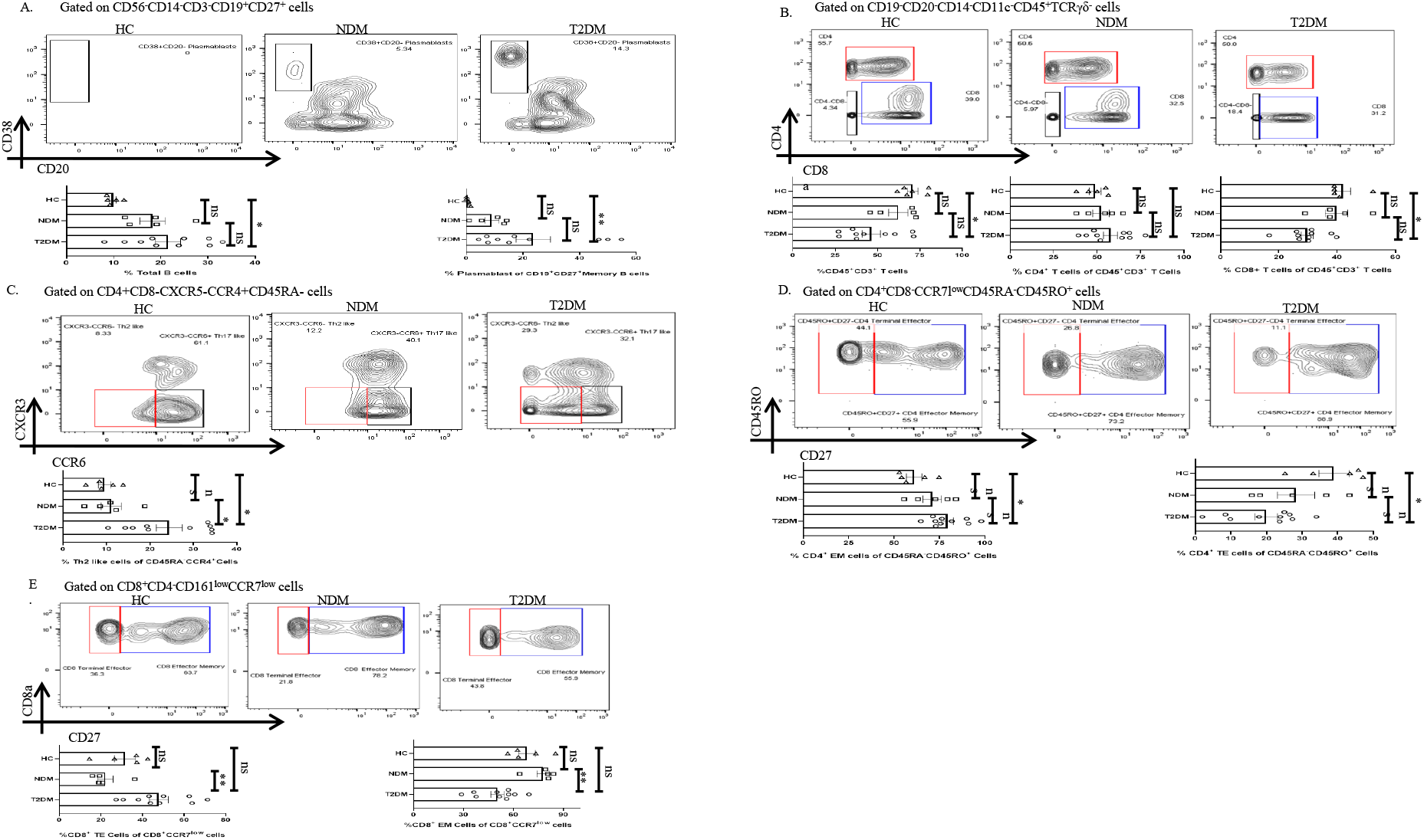
Adaptive Immune Cells in T2DM and NDM patients. Whole blood from Covid-19 positive T2DM(n=10), NDM(n=5) and Covid-19 negative and non-diabetic healthy controls (n=5)was stained heavy metal tagged antibody and analyzed in mass cytometer.**(A)**Representative Gating strategy to identify CD38^+^CD20^-^ Plasmasblasts. Statistical analysis of percentage of plasmasblasts. **(B)** Gating strategy to identify CD45^+^CD3^+^CD4^+^ and CD45^+^CD3^+^CD8^+^ T cells. Statistical analysis for the frequency of CD3^+^,CD4^+^ and CD8^+^ T cells.**(C**) Gating strategy for CD4^+^CXCR3^-^CCR6^-^Th2 like cells.Statistical analysis for the percentage of Th2 like cells.. **(D)** Gating strategy to identify CD4^+^ TE(CD45RO^+^CD27^-^)and CD4^+^ EM cells(CD45RO+CD27^+^). Statistical analysis for the percentages of CD4^+^ TE and EM cells.**(E)** Gating strategy to identify CD8^+^ TE (CCR7^-^ CD27^-^) and EM(CCR7^-^CD27^+^) cells. Statistical analysis for the percentages of the CD8^+^TE and EM cellsThe Kruskal Wallis Test (non parametric) with post hoc Dunn’s multiple comparison test was performed p<0.05 was considered statistically significant (*), p<0.01 was considered to be very significant (**), *P* < 0.001 was considered to highly significant(***), *P* < 0.0001 was considered extremely significant(****) ns, not significant. All error bars were SEM.

### High Dimensional analysis reveals immune perturbations in T2DM patients

We also performed t-distribution stochastic neighbour embedding (t-SNE) analysis to understand how different markers reported previously, show variability in Covid-19 patients in our population of study. t-SNE analysis revealed a decrease in the expression of CD3 and CD8 in T2DM patients as compared to healthy volunteers **(Fig. 6A)**. On the contrary, there was increase in the expression of CD14, CD38 and HLA-DR in T2DM patients as compared to healthy volunteers **(Fig. 6B)**.

**Fig. 6.**
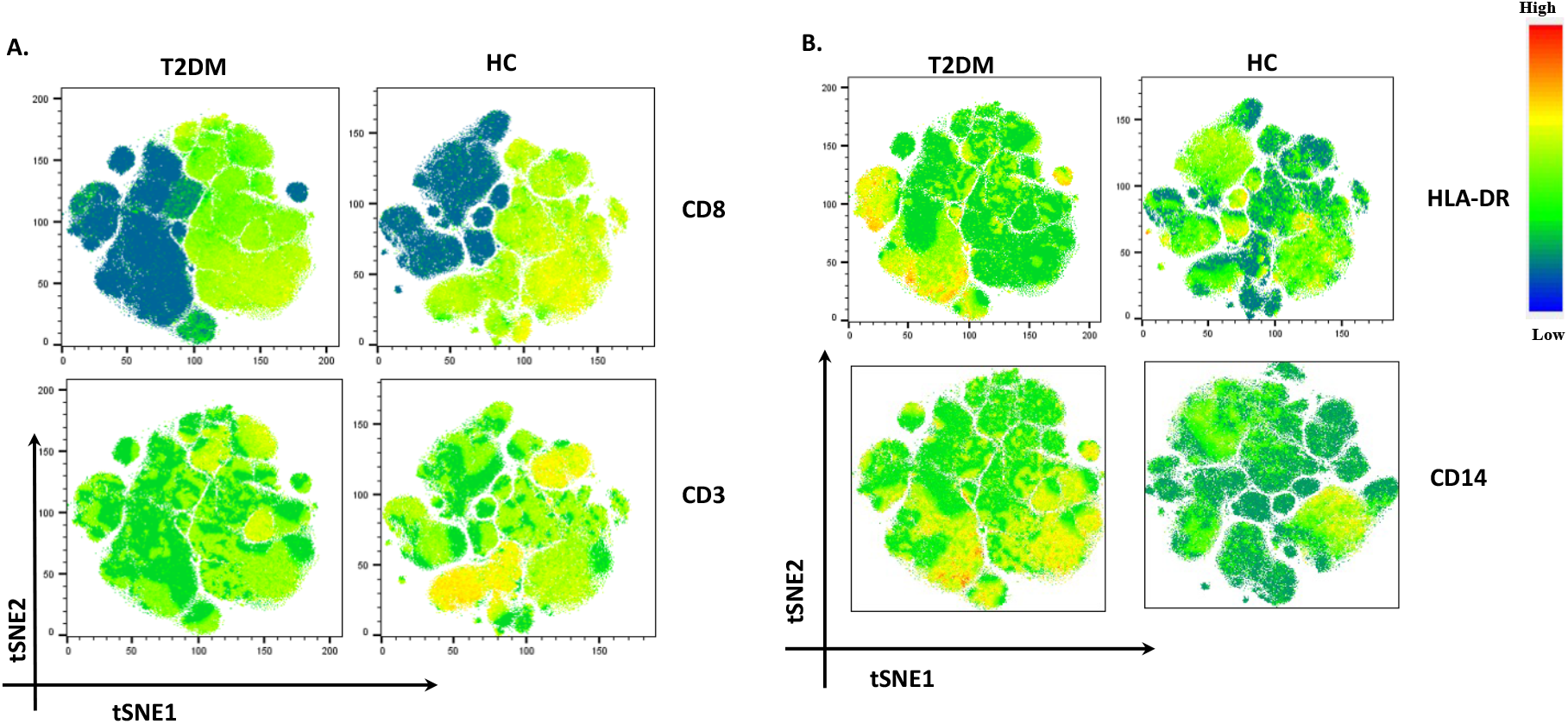
t-SNE analysis of lowest and highest expressed markers in T2DM patients. **(A)** CD8 and CD3 showing decreased expression in T2DM patients. **(B)** HLA-DR and CD 14 showing increased expression in T2DM patients when compared to controls.

## Discussion

The Covid-19 pandemic has brought an unprecedented global devastation on lives and livelihood of the common people, breakdown of the best healthcare system and economic collapse all across the globe. Till October 2021, India reported around 34 million cases, being second in place to the United States of America. Unfortunately, a bulk population of Covid-19 cases in India who had succumbed to the infection had comorbidities and amongst these, Type II Diabetes was the most common. At present, there are no significant reports from India providing an insight into the variations in immune response, extent of inflammation and outcome in T2DM patients as compared to NDM patients and healthy volunteers. In this study, we attempted to describe and compare immune response, viral loads and clinical parameters amongst T2DM, NDM and healthy volunteers. In that respect, 25 patients with T2DM, 10 NDM patients and 10 healthy volunteers were studied for cytokine, chemokine, viral loads, clinical parameters and antibody isotyping assays for understanding the immunopathology of the virus. Ten T2DM, five NDM and five healthy volunteers were studied for whole blood mass cytometry assay to determine the difference in immune cells amongst the groups. While it is understood that this is a very small group, some clear patterns are established here suggesting that the chronic Meta inflammation in Type II Diabetes might play a role in further precipitating the inflammatory signatures in Covid-19.

It is known that mild inflammation serves as one of the major initiator mechanisms of the immune system that is extremely crucial in fighting infection (14). However elevated levels of circulatory cytokines and eventual cytokine storm and dysregulated inflammatory responses have been evinced in Covid-19 infection (2, 4). Additionally, previous reports suggest that patients with T2DM have a higher vulnerability for SARS-CoV2 infection (15, 16) where persistent low grade inflammation associated with T2DM was evinced but not co related to be causal. Our data taken together with biochemical, cytokine and antibody profile strongly indicate that chronic low grade inflammation and the consequential dysregulated immune response might be responsible for the upsurge of inflammatory responses in Covid-19 patients leading to severe infection and or mortality.

Our preliminary objective in the study was to assess the differences in viremia in Oropharyngeal swab between the two groups. However, viral load of OP samples did not show any significant difference between the two groups. This was in accordance with previous studies, where it was observed that the viral load in nasal and throat swab samples were similar in the NDM and T2DM patients (17, 18). Here, the T2DM are considered as severe Covid-19 patients, based on the severity of their symptoms. And, the NDM patients either had NDM symptoms (n=4) or were asymptomatic (n=6). This is evident from the clinical parameters as T2DM patients had higher levels of CRP which is a non-specific inflammatory marker, dysregulated glucose metabolism as evident from higher plasma glucose, glycated haemoglobin and impaired blood coagulation as interpreted from higher d-dimer levels(19-22).

Cytokines and chemokines are soluble mediators that control the migration of immune cells to the site of inflammation, providing a pro or anti-inflammatory environment which essentially shapes the type of immune response during a disease. In our T2DM patients who were infected with SARS-CoV2, higher expression of both type I and type II cytokines were evinced. Previous reports suggest the involvement of both the types of cytokines in aggravating the severity of the disease. Increase in type I cytokines such as IFN-γ TNF-α, IFN-α2, IL-6, GMCSF, IL-8 indicated a robust antiviral and inflammatory response occurring in these T2DM patients. Additionally, these patients also showed an increase in type II cytokines such as IL-4 and IL-9, representing the anti-inflammatory response. Here, we understand that the presence of both inflammatory and anti-inflammatory cytokines in T2DM patients as indicators of a dysregulated immune response where the anti-inflammatory response was clearly incapable of shutting the cytokine storm in these patients(23).

In diabetic patients, there is an increase in the inflammatory cytokines, CRP levels and the presence of blood clots. As similar symptoms are associated with Covid-19 i.e. elevated CRP levels, D-dimer and inflammatory cytokines, this suggests towards a synergistic effect found in T2DM patients infected with SARS-Cov2, which was reduced in NDM patients. Additionally, our study suggested an increase in IL-15 and 1L-7 in T2DM patients when compared to NDM and healthy volunteers. As it is known that IL-15 is crucial for promoting cytotoxic activity of both, NK and CD8^+^ T cells and is also involved in memory CD8^+^ T cell differentiation, it should have led to increase in the CD8^+^ population(24). However, we found a decrease in the CD8^+^ cell population, specifically in the EM compartment, again indicating a dysregulated or non-responsive immune response in these patients. This might be a consequence of reduced receptors for IL-15 in these cells. As IL-7 is also required for T cell development and maintenance(25), its increased numbers in T2DM patients partially corroborated with the increase in CD8^+^ TE cells and CD4^+^ EM cells. However, as a consequence of immune dysregulation, neither the elevated levels of the above mentioned cytokines nor the CD8^+^ T cells, are capable of controlling the inflammation.

The sCD40L is one of the major co-stimulatory molecules on activated T cells that interact with CD40 on B cells and is responsible for immunoglobulin isotype switching in the membrane-bound form(26). In our study, levels of sCD40L were elevated in NDM as compared to the T2DM patients indicating that the NDM patients may be undergoing an immuno suppressive reaction, as elevated sCD40L is also associated to immunosuppression. GRO (CXCL1) is a chemokine that attracts variety of immune cells, particularly neutrophils and is also implicated in wound healing process(27). Similar to sCD40L levels, GRO levels were also higher in NDM as compared to T2DM patients indicating that both inflammation and healing were occurring simultaneously in these patients. However, the healing mechanism was definitely poorer in T2DM patients. The levels of Macrophage-derived chemokine (MDC/CCL2) were also higher in NDM patients than T2DM patients. As MDC is known to be elevated in lung inflammation and hemorrhage but reduced in T2DM symptomatic patients, this is also suggesting towards a possible immunosuppressive function of MDC in this population(28).

To understand how different antibody isotypes influence the clinical phenotype in T2DM and NDM patients, we did an antibody isotyping from the plasma of these patients. Here, we found significant difference in the levels of IgA, IgM, IgG1, IgG2 and IgE between the two groups but IgG3 and IgG4 did not show any significant difference and we understand that the structure and function of different antibody isotypes vary from one another. For example, IgG1, IgG2 and IgG3 can fix complement(29). However, IgG4 cannot fix complement and is unable to induce antibody mediated cell cytotoxicity (ADCC). Similar to previous reports, we found an increase in the levels of IgE in T2DM patients(8, 16). This corroborated with the increase in the Th2 subset along with increase in cytokines, IL-4 and IL-9 in T2DM patients, as it is known that IgE is associated with Type II response. In addition, there was also an increase in IgG1 and IgG2 in these patients, indicative of the inflammatory response. However, as IgE was high in these T2DM patients, we could see no significant difference in IgG4 levels. This could be accredited to the competition between IgE and IgG4 against each other, for fixation sites in basophils and mast cells, as suggested by previous reports(30). IgA levels were also found to be higher in T2DM, indicating towards viral immune response at mucosal surfaces(31). With respect to IgM, its increased levels indicate towards an ongoing infection. However, the lack of any longitudinal profiling restricts us from understanding its specificity to antigen(32).

With respect to our whole blood analysis by mass cytometry, we found an increase in the percentage of granulocytes in T2DM patients when compared to healthy controls indicating underlying inflammatory responses in these patients. This is consistent with other reports which suggested an increase in granulocytes with increasing severity(15, 33). On the other hand, we found a decreasing percentage of monocytes in T2DM patients as compared to NDM patients **(Table 1)**, although the difference was not significant. This has also been reported in a previous study showing the absence of any significant changes in the total monocytes and its subsets in T2DM patients with various comorbidities, T2DM being one of them(15). Additionally, we also found that T2DM patients showed a decreasing trend in lymphocyte percentage as compared to NDM and healthy volunteers, as reported in previous studies (4, 8, 15). One of the significant findings from our study is that, there is a drastic reduction in the percentage of total dendritic cells in T2DM patients when compared to both, NDM and healthy controls. Within the dendritic cell compartments, there is a decrease of pDCs but mDCs have increased in T2DM patients when compared with healthy controls. In general, pDCs are known to secrete type I interferons in response to viral infection(34). However, our study has shown a decrease in the percentage of pDCs, but increase in the cytokine IFN-α2. This is suggesting that although these cells are secreting high levels of type I interferon, their numbers are depleted in T2DM patients and thus, are unable to control the infection. With respect to mDCs, they are known to secrete the cytokines IL-12 and TNF-α, which polarizes the T cell towards a type I response, crucial for controlling the viruses(35, 36). Our study also observed an increase in the population of mDCs along with increase in the cytokines, IL-12p40, TNF-α in T2DM. Although certain reports have suggested a decrease of mDCs along with pDCs, we observed the presence of high numbers of mDCs in diabetic patients, indicating an ongoing low grade inflammation, which is further augmented with Covid-19 infection. However, since we did not do a longitudinal profiling, we could not delineate the mDC dynamics in these patients(37, 38).

With respect to basophils, literature suggested a decrease in its population with increasing severity(39). We found a similar trend, as the basophil population decreased in T2DM patients as compared to healthy controls. Similarly, MAIT/iNKT cells were significantly decreased in T2DM patients as compared to NDM and healthy controls. However, no significant differences were observed in the percentage of gamma delta T cells. A decrease in MAIT/iNKT could indicate towards a migration to other inflamed tissues including the lungs, as suggested by the previous reports. With respect to NK cell population, it is known that they play a crucial role in fighting viral infections and previous reports suggested a decrease in their numbers in T2DM cases. However, our population of study did not show any significant difference amongst the groups and as mentioned earlier may be a consequence of the time of sampling of patients(40, 41).

In case of B cells, we observed a slight elevation in its percentage in T2DM patients, when compared to NDM and healthy controls, but statistical difference was found only between T2DM patients and healthy volunteers. Although no difference was observed in naive or memory B cell compartment, plasmablasts were significantly elevated in T2DM patients as compared to healthy controls. The increase in plasmablasts in T2DM Covid-19 has also been reported in other studies (8, 15)but unlike our study, they could not correlate it with any comorbidity. Interestingly, there are reports suggesting an increase in extrafollicular B cell response in Covid-19 and other inflammatory diseases such as Systemic Lupus Erythematous. Since most of the Covid-19 patients had type II diabetes in our study, we hypothesize that these two factors might have augmented the plasmablast production. Although we did not analyze the clonality of B cells, but earlier study has suggested an oligoclonal expansion of B cells in T2DM Covid-19, in turn correlating with increased plasmablast production. Increased B cell population in T2DM patients also corroborated with the fact that most of the immunoglobulins (IgM, IgA, IgG and IgE) were elevated in these patients as opposed to NDM and healthy volunteers. Altogether, we found an increase in the plasmablast population in T2DM as compared to NDM patients and healthy volunteers, suggesting that the low grade inflammation in Diabetic patients resulted in increase in plasmablast population, which augmented further with Covid-19 infection in these patients.

T2DM patients also displayed a lower percentage of CD45^+^CD3^+^ T cells as compared to healthy controls. Within CD3^+^ T cell population, there was a decrease in the percentage of CD8^+^ T cells in T2DM as compared to healthy volunteers. However, CD4^+^ T cells did not show any statistically significant difference amongst the groups. Earlier reports have suggested the loss of CD8^+^ T cells being greater than CD4^+^ T cells. Since our population of study was small with heterogenous manifestation, this could explain why CD4^+^ T cells did not show any difference amongst the groups. Additionally, several reports indicate towards T cell apoptosis or migration to other tissues in SAR-CoV2 infection, leading to decrease in T cell population in the periphery and could account for the decrease in T cells in the T2DM patients(42-44). In general, there was decrease in the CD3^+^ T cell population, where the CD8^+^ compartment was affected more significantly than CD4^+^.

Type II diabetes has been reported to have various aberrancies such as an impaired differential potential and secretion of multiple proinflammatory cytokines such as TNF-α and IFN-γ(37, 45). Although we could not find any significant difference amongst the groups in the percentage of Th1 like, Th17 like, Treg or cTfh subtypes, there was an increase in the proinflammatory cytokines. We understand that the absence of difference amongst the groups with respect to T cell subsets can be accounted to the time of sampling. On the other hand, our study found an increase in the percentage of Th2 cells secreting type II cytokines such as IL-4 and IL-9. The high numbers of Th2 cells partially explained the augmented levels of immunoglobulins in these patients, specifically IgE which is induced by cytokines such as IL-4. Taken together, this indicated that the Diabetic patients already had low grade inflammation. When encountered with Covid-19, dysregulation in the immune system escalated to such levels that even with the increase in Th2 population secreting anti-inflammatory cytokines, inflammation could not subdue.

We also analyzed for the naïve, effector and memory compartments in both CD4^+^ and CD8^+^ T cells. In CD4^+^ T cells, we did not see any difference in the overall percentage of naive (CCR7^+^CD45RA^+^CD45RO^-^) and central memory cells (CCR7^+^CD45RA^-^CD45RO^+)^ amongst the various groups. However, effector memory cells (CCR7^-^CD45RA^-^CD45RO^+^CD27^+^) were elevated in T2DM patients when compared to healthy controls, suggesting cross reactivity with other family of coronavirus, as reported by a previous study from India(46, 47). An important point to note here is that central memory cells did not show any significant difference and this may be due to the fact that CM cells are lymph node residents as opposed to EM cells, which circulate in blood and thus, respond faster to the antigen (data not shown). However, since we did not analyze for the antigen specific cells, therefore we were unable to find any significant difference in the percentage of CM cells. Terminal effector (CCR7^-^CD45RA^-^CD45RO^+^CD27^-^) cells were decreased in T2DM patients with Covid-19 as compared to healthy controls, corroborating with the observation that these cells are first to react with the virus and the subsequent proinflammatory cytokine milieu is responsible for the apoptosis of these cells.

In CD8^+^ T cell compartment, EM (CCR7^-^CD27^+^) cells were elevated in NDM as compared to T2DM patients, indicating an effective immune response in these patients but less effective in T2DM patients. TE (CCR7^-^CD27^-^) cells increased in T2DM patients indicating that these cells are highly inflammatory and might be insensitive to AICD (47-49).

Altogether, our data suggests that Meta inflammation present in these T2DM patients is responsible for aggravation of the inflammation occurring due to Covid-19. With T2DM being one of the most common morbidity presents in India, these patients remain most vulnerable and were susceptible to secondary infections like mucormycosis. Though vaccinations are lowering the severity of the disease, reported waning antibody response in 3-6 months is a major concern apart from how long the protective immunity from T cells will work in these patients is also yet to be ascertained. Our study is essentially aimed at understanding how a low grade chronic inflammatory disorder such as Type II Diabetes dictates the immune response during the pathogenesis of Covid-19. We understand that our study is not devoid of limitations such as a small sample size, absence of longitudinal study or follow up, but nevertheless it will help in understanding the vulnerability of these patients and subsequent planning of vaccine coverage in these patients or future vaccines booster dosages to them.

